# Chest binding practices and health impacts for transmasculine individuals: A mixed methods analysis

**DOI:** 10.1101/2025.06.14.25327031

**Authors:** Theo G Beltran, Ashleigh J Rich, Mannat Malik, Erin Cooney, Jean-Michel Brevelle, Katherine Croft, Rachel Bluebond-Langner, Zach Reilly, Tonia Poteat

## Abstract

**Background:** There is a large knowledge gap among health care providers regarding the importance of chest-binding among transmasculine people. With increased passage of anti-transgender laws denying rights and health care services for transgender youth, it is more important than ever to document their experiences within healthcare settings.

**Methods:** This mixed methods study integrated quantitative and qualitative data. Participants were transmasculine individuals who completed surveys (N=44) or in-depth interviews (N=21) in Baltimore or Towson metropolitan areas in 2016.

**Results:** The majority of transmasculine survey participants bound in their lifetime (n=36, 82%). Among them, 43% (n=19) reported binding seven days per week and 52% (n=23) bound eight or more hours per day on average. Of survey participants, 39% felt healthcare providers were not comfortable with transgender people. Interview themes included physical health challenges of binding, provider de-prioritization of binding, gender affirmation as a facilitator of safety, need for safer binding education, and binding as mental health promotion.

**Conclusion:** Binding is an important part of gender affirmation for many transmasculine people, with social, safety and mental health benefits. However, many transmasculine people report barriers to social and healthcare provider support around binding. Healthcare providers should understand the benefits of binding and be prepared to discuss safer binding practices with transmasculine patients.

## Introduction

Chest dysphoria (the experience of gender-related distress caused by having breasts) is experienced by many gender diverse youth and adults and can lead to adverse mental health impacts.^1^ In a study of transgender and gender diverse youth aged 13 to 24 years, chest dysphoria was significantly associated with poor life satisfaction.^2^ Chest binding refers to the compression of breast tissue to create a flatter or more masculine appearing chest.^3^ Binding is a necessary practice for many transmasculine (the gender identity of a person who is partially or fully masculine, which differs from their sex assigned at birth) people to better align with their gender expression and participate in everyday life.^3,4^

However, transmasculine people may not bind due to overheating or discomfort while binding, or financial barriers to binders.^5–7^ Transmasculine people interested in chest surgery may use binding as an interim method before surgery.^8^ Chest surgery, which is the removal of chest tissue to obtain a flatter chest that may be perceived as masculine, has shown to alleviate chest dysphoria and mental health distress.^1,9^ With parents, guardians, state policies, and insurers often limiting access to gender-affirming surgeries, in addition to financial and distance barriers, chest surgery is inaccessible to many.^10^

Chest binding has been linked to improved social confidence and reduced anxiety, depression, and suicidality.^7^ In-depth interviews in Australia found that while binding improved self-reported mood, frequent binding was associated with physical discomfort, such as not being able to breathe deeply.^4,7^ Still, participants described binding as distress relieving, and affirmed prioritizing their mental health over physical comfort.^4,7^

Evidence of low competency among healthcare providers regarding binding practices has been reported. In the Binding Health project, 60% of participants reported their healthcare provider reacted negatively to their binding.^7^ In a cross-sectional study evaluating binding impacts for gender-diverse adolescents, only 3% reported learning of binding from providers and nearly all participants taught themselves through online sources.^2^ This highlights a large knowledge gap among providers.

With increased passage of anti-transgender laws denying rights and health care services for transgender youth, it is more important than ever to document their experiences within healthcare settings. Qualitative context is needed to support quantitative data of binding experiences to understand binding practices and inform recommendations. This mixed methods analysis aims to explore chest binding practices and health impacts for transmasculine people to inform transgender-competent clinical practice.

## Methods

### Study Design and Population

This mixed methods analysis integrated descriptive statistics from a quantitative survey, which informed thematic analyses from qualitative in-depth interviews.^11,12^ Quantitative survey data were drawn from the Be the Conversation (BTC) study. Informed by formative research with local stakeholders, BTC aimed to explore the health and social needs of transgender Baltimoreans.

Surveys were conducted between March and September 2016. Qualitative data was drawn from the TOP study between July and November 2016, which aimed to explore transgender men’s surgical goals and post-operative experiences related to chest surgery. Protocols and procedures for both studies were approved by the Johns Hopkins School of Public Health Institutional Review Board.

### Eligibility

Individuals were eligible for the BTC study if they were (1) age ≥18 years, (2) lived in the Baltimore or Towson metropolitan areas, and (3) identified as transgender or any gender different from their sex-assigned-at-birth. For this analysis, data was limited to BTC participants who were assigned female sex-at-birth (N=44), referred to hereafter as transmasculine individuals. Participants reviewed an online consent form and consented prior to participation.

In the TOP study, 9 transmasculine individuals seeking chest surgery and 12 who had undergone chest surgery completed in-depth interviews (total=21). Individuals were eligible to participate if they were: (1) age ≥18 years, (2) assigned female sex-at-birth and identify as male/man or transgender, and (3) had or planned to have chest surgery. TOP participants provided verbal informed consent.

### Recruitment and Data Collection

#### Quantitative

Participants were recruited using multiple convenience methods, including partnering with trusted community organizations to distribute study flyers, outreach at local Lesbian, Gay, Bisexual, Transgender and Queer (LGBTQ) community events, social media engagement, and word-of-mouth. The survey was self-administered online through the SurveyMonkey platform. In-person surveys were interviewer-administered in a private space to accommodate a range of literacy levels. Participants received a $25 Visa gift card.

#### Qualitative

Participants were recruited at two surgical clinics offering chest surgery. Partnerships were established with clinic providers, who referred eligible patients. Other participants were referred by local LGBTQ organizations. Interviews were conducted by three cisgender study members in private rooms at the clinics or online and ranged from 32 minutes to 2 hours (average 55 minutes). All interviews were recorded and professionally transcribed.

### Quantitative Measures

The BTC survey included questions about chest binding experiences adapted from the Binding Health Project survey.^7^ Binding was defined in the survey as a “non-surgical method to reduce the size of the chest to be more comfortable with one’s gender expression.” Transmasculine participants were asked if they ever bound, were currently binding, their binding practices (current practices or former for individuals who have undergone chest surgery), concern about potential health impacts of binding, and health problems potentially caused by binding. Participants received a list of health concerns they attributed to binding.

### Data Analysis

#### Quantitative

Quantitative data were analyzed using R Studio (PBC, Version 4.1.2). Descriptive statistics on binding practices and experiences with healthcare providers are reported. Results by binding history are presented to explore differences in practices and needs of participants.

#### Qualitative

The semi-structured interview guide covered five domains: (1) gender identity and expression, (2) motivations for seeking chest surgery, (3) experiences accessing chest surgery, (4) surgical goals and concerns, and (5) experiences following surgery. An *a priori* codebook was developed based on the interview questions and revised to include emergent thematic codes based on transcript reviews and team discussion. “Chest binding experiences” was included as a thematic code. The interviewers conducted side-by-side coding for three transcripts. Discrepancies were resolved through consensus. The study team reviewed all chest binding experiences quotes and selected salient themes collaboratively.

## Results

### Sample Characteristics

The average age was 32 years while the majority of transmasculine participants were White (n=31, 71%), completed a college degree or higher (n=27, 61%) and had private health insurance (n=25, 58%) (**Table 1**). Half of transmasculine participants were living on an income below the federal poverty line (n=22, 50%) and had food insecurity in the past 30 days (n=13, 30%).

**Table 1.** Socio-demographics of transmasculine survey participants.

| Sample Characteristics | Total<br>(N=44)<br>n (%) | Ever Bound<br>(n=36) | Never Bound<br>(n=7) |
| --- | --- | --- | --- |
| Mean age (Range) | 32 (19-54) | 32 (19-53) | 36 (24-54) |
| Mean age initiated binding (Range) | 28 (17-51) | 28 (17-51) | NA |
| Race/ethnicity |  |  |  |
| White | 31 (71) | 26 (72) | 4 (57) |
| Black or African American | 4 (9) | 4 (11) | 0 (0.0) |
| Multi-racial | 5 (11) | 5 (14) | 0 (0.0) |
| Latinx/Hispanic (any race) | 5 (11) | 4 (11) | 1 (14) |
| Highest level of completed education |  |  |  |
| High school degree or less | 17 (39) | 12 (33) | 5 (71) |
| Some college or greater | 27 (61) | 24 (67) | 2 (29) |
| Health insurance |  |  |  |
| Private | 25 (58) | 22 (61) | 3 (43) |
| Public | 16 (37) | 12 (33) | 4 (57) |
| Uninsured | 2 (5) | 2 (6) | 0 (0.0) |
| Socioeconomic factors |  |  |  |
| Unstably housed, past 12 months | 9 (21) | 8 (22) | 1 (14) |
| Income below federal poverty line | 22 (50) | 17 (47) | 4 (57) |
| Unemployed | 8 (18) | 6 (17) | 1 (14) |
| Any food insecurity, past 30 days | 13 (30) | 11 (31) | 2 (29) |
| Gender affirming care |  |  |  |
| Tried to access gender-transition related care in past 12 months | 31 (72) | 29 (81) | 2 (29) |
| Tried to access care and unable to, past 12 months | 5 (11) | 5 (11) | 0 (0.0) |
| Ever taken hormones | 28 (64) | 25 (69) | 3 (43) |
| Currently on hormones (among ever taken hormones) | 24 (86) | 22 (88) | 2 (67) |
| Had top surgery | 17 (39) | 15 (42) | 2 (29) |
| Experiences of Discrimination |  |  |  |
| Provider not comfortable with transgender patients | 17 (39) | 15 (42) | 2 (29) |
| Person treating them tried to stop them from being trans | 11 (25) | 10 (28) | 1 (14) |
| Parent or caregiver tried to stop them from being trans | 10 (30) | 9 (25) | 1 (14) |
| Healthcare provider disrespected them and their friends in the past | 8 (18) | 6 (17) | 2 (29) |
| Binding history |  |  |  |
| Lifetime | 36 (82) | 36 (82) | NA |
| Currently | 13 (30) | 13 (30) | NA |
| Mental Health |  |  |  |
| PTSD (modified PC-PTSD-5) | 14 (32) | 12 (33) | 1 (14) |
| Depression (PHQ-2) | 15 (34) | 13 (36) | 1 (14) |
| Anxiety (GAD-7) | 18 (41) | 16 (44) | 1 (14) |

Abbreviations: N= Number; NA= Not Applicable; PTSD= Post-traumatic Stress Disorder; PC-PTSD-5= Primary Care Post Traumatic Stress Disorder Screen for Diagnostic Statistics Manual 5; PHQ-2= Patient Health Questionnaire-2; GAD-7= General Anxiety Disorder-7.

Over half (n=28; 64%) of participants had ever taken hormones and among them, 86% (n=24) were currently on hormones. There were no major differences between participants with binding history and those without **(Table 1)**.

Characteristics of the twenty-one qualitative interview participants are described in **Table 2**. The average age was 31 years and participants were mostly White (n=14, 67%). Five qualitative themes are described below in connection to quantitative results. A joint-display table of the convergence (agree), divergence (disagree), or expansive relationship (provides new insight) between quantitative and qualitative results are in **Table 3**, with example quotes in **Table 4**.^13,14^ The joint-display table results were decided upon based on whether qualitative and quantitative results aligned, did not align, or there was new insight offered from either.^15^

**Table 2.** Socio-demographics of qualitative interview participants.

| Sample Characteristics | Total (N=21) | Transgender Men Pre-Chest Surgery (n=9) | Transgender Men Post-Chest Surgery (n=12) |
| --- | --- | --- | --- |
|  | n (%) | n (%) | n (%) |
| Mean age (Range) | 31 (20-49) | 34 (20-49)<sup>a</sup> | 28 (21-42) |
| Race/ethnicity |  |  |  |
| White | 14 (67) | 5 (56) | 9 (75) |
| Black or African American | 4 (19) | 3 (33) | 1 (8) |
| Multi-racial | 3 (14) | 1 (11) | 2 (17) |

Abbreviations: N=Number
<sup>a</sup>Age missing for one participant—participant is over 23 years based on age information reported during interview.

**Table 3.** Joint display of transmasculine experiences of binding.

| **Finding** | **Quantitative Results** | **Qualitative Results** | **Converge, Diverge, Expand** |
| --- | --- | --- | --- |
| Theme: Physical challenges of binding |  |  |  |
| Binding was associated with pain and other physical challenges | Transmasculine participants who bound attributed health concerns to binding including back pain, shortness of breath, bad posture, and shoulder pain. | Transmasculine participants who bound reported high levels of physical pain. | Convergent |
| Physical challenges of binding impacted essential daily activities | The majority of transmasculine participants who bound expressed concern about the effects of binding on their health. | Transmasculine participants worried about the physical challenges of binding and their impact on ability to take part in daily activities like work and sleep. | Convergent |
| Binding materials were major contributors to physical health impacts | More than half of those who bound using binders, sports bras, or shirt layering experienced back pain. | Depending on the binding material, participants experienced physical effects such as a full body itch. | Expand |
| Theme: Provider and caregiver de-prioritization of binding |  |  |  |
| Provider and caregivers de-prioritized binding and devalued its role in gender affirmation | Transmasculine survey participants reported discrimination by healthcare providers, related to transgender health and specifically binding. | Participants experienced providers and other caregivers insisting on stopping binding, and not recognizing it as essential for their health. | Convergent |
| Providers lacked experience and sensitivity in gender-affirming care and making informed recommendations around binding | Nearly half of participants felt that healthcare providers were not comfortable with transgender people and a quarter had a healthcare professional try to stop them from being transgender. | Transmasculine participants encountered insensitivity and inexperience from health care providers and caregivers regarding binding. | Convergent |
| Barriers to receiving optimal care and support around binding included provider discrimination and cost. | Experiencing disrespect from healthcare providers was a barrier to accessing care. | Financial costs were a barrier to accessing binding materials. | Convergent |
| Theme: Gender affirmation as a facilitator of safety |  |  |  |
| Binding was an important element of gender affirmation, serving as a facilitator of safety for participants | Almost all participants noted that being perceived as cisgender in public was important. | Binding served as a form of protection from interpersonal discrimination and led to feelings of safety. | Convergent |
| Social affirmation and binding | Over half of binding participants had a high score on the gender identity non-affirmation scale. | Participants felt that binding helped them from being visibly non-cisgender passing and experiencing microaggressions and discrimination. | Divergent |
| Theme: The need for safer binding practices |  |  |  |
| Participants emphasized the need for safer binding practice education | Binding was a common experience for transmasculine participants. Almost all survey participants had experience with binding, either currently or in the past. | Transmasculine participants discussed the urgent need for safer binding practice education, especially when they began binding. | Expand |
| Safer binding materials were not accessible to participants | Participants who used ace bandages or duct tape for binding named financial concerns as a barrier to accessing health care and the length of time they bound was from 1 to over 7 years. | Participants bound for long periods of time and didn't have access to safe binders, which led to long term health impacts. | Expand |
| Many participants bound outside clinical recommendations | When binding, nearly half reported binding 7 days per week and just over half bound 8+ hours per day on average. Most participants bound for 1 to 3 years total, suggesting that long-term binding is a common practice | Participants bound all day and every day of the week. | Convergent |
| Binding promotes better mental health |  |  |  |
| Mental health as a reason for choosing to bind | Participants reported several poor mental health outcomes, including PTSD, and anxiety and depression symptoms. | Binding was centrally important to supporting the mental health of transmasculine participants. | Expand |
| Binding and its impact on mental health | There were no major differences in prevalence of mental health conditions by binding history, although sample size is too small to test statistical significance | Binding helped build self-esteem and made participants feel 'normal' as transmasculine people | Expand |

**Table 4.** Qualitative Themes and Example Quotes.

| Themes | Example Quotes |  |  |
| --- | --- | --- | --- |
| Physical challenges of binding | “I’m also a waiter, so like all of that movement and just nonstop not being able to sit down and...Or anything like that, on top of wearing a binder all day, like...After a while it built up in my fibro, which is...It was really bad. I could sleep, but during the day it just...Like there were at least a few times that I had breakdowns at work from the pain, and they had to send me home.” | “I had a lot of issues with - kind of like right here - I don’t want to say it was my esophagus but eating was hard to do, but mostly just my back and the full body itch.” | “I binded for about six years and because it compresses everything it puts a lot of tension on your back and whenever I’d take it off the first thing I had to do was have to lie down flat on my back and just kind of like re-stretch my back. Then when it would get too hot out it was just like a full body itch because the material is not very breathable. Binding was one of the - it was a necessity but it was one of the worst things I had to do.” |
| Provider and caregiver de-prioritization of binding | “She said, ‘You’ve got to take it off.’ And I just broke down and cried to her because I was like, ‘I can’t, I can’t take it off.’ I don’t care if it were to kill me. To me it would kill me to take it off. So, unfortunately, that’s how our life is.” | “It hurts and it’s painful and, on the other hand, it’s that my back, I have a bad back. My back doctor told me that I can’t wear the binder anymore. So, with all that other stuff, it’s like, ‘Well I am going to have to wear the binder.’ So, she’s like, ‘Well isn’t there a surgery that you can get so you don’t have to wear the binder?’ And I’m like, ‘Yeah, but it’s expensive.’ And so she was like, ‘Well, try to get that surgery because it’s hurting your back more. Your binder’s hurting your back.’ So that’s another reason why personally I’ve looked for a surgery.” | “Binding, for me, has been an issue due to cost; how much the material costs. You want to get the best based off of what others may recommend, who makes it; and then the different results you would get from wherever you get it from. So I’ve gone some cheap routes because of that. I don’t have \$60, \$70, \$80 to get the different types of pieces that will give you my best look and not rise up on me and not compress my ribs, not give me a pushup – all these different factors. It’s too much.” |
| Gender affirmation as a facilitator of safety | “I think the most part is that technically it has nothing to do with anybody else, but it has everything to do with everybody else at the same time. Because, personally, if I take it off then I am worried about what everybody else would think because society doesn’t see a male have boobs. And in society’s eyes I look like a male now. And I mean, technically, all that they could say is be mean to me and ridicule me, but nobody wants that. And what the binder does is hide that and make me seem quote, quote normal and also it can be like a safety issue.” | “Yeah; I think it’s just - for me at least and I think for a lot of my friends - it’s one of the most important steps because it’s affirming, both for that individual and then also for people outside looking in, because then it’s one last thing that people have to stop and be like, “What’s that? Why do you have man boobs?” |  |
| The need for safe binding practices | “Well, when I was first coming out as a trans person, I used an Ace bandage. Obviously that’s a lot of people’s first go-to. Until I learned firsthand how dangerous they can be. Wearing it on a hot day, tight as possible, it hurts your ribs, it really does. And I...Like I said, I learned that the hard way. I almost passed out, I couldn’t breathe, my ribcage felt like it was pressing in. And, obviously, if I had known that there was such a thing as a binder, a proper binder that won’t hurt you, I probably would’ve used it. But at that time, I was a kid. The first thing that I saw on the internet was a guy wearing an Ace bandage. Oh, that makes sense. It’s skin colored, you know, and it binds your chest, that’s fine. So I wore it, not knowing how dangerous it could be.” | “I started binding probably once I started growing a chest. I don’t know, maybe I hit puberty, so 12, 11... So I started duct taping myself, which was horrible because it ended up ripping off my skin really bad. I didn’t care... So then I found out about binders, probably when I was like 18. But at this time, my back and my body was shot.” | “I wear them pretty much all day every day, until I go to sleep, that is.” |
| Binding promotes better mental health | "...a lot of people, including myself, would wear it just for the full day just because it makes you seem - gives the impression of flat chest so it helps passability and comfortability even if you’re not physically comfortable with the binder on. It gives you almost like that mental peace of mind. I binded for about six years and because it compresses everything it puts a lot of tension on your back and whenever I’d take it off the first thing I had to do was have to lie down flat on my back and just kind of like re-stretch my back. Then when it would get too hot out it was just like a full body itch because the material is not very breathable. Binding was one of the - it was a necessity but it was one of the worst things I had to do.” | “I mean I would just probably say my mental health, which is hugely important because our thoughts become our actions. So just feeling so much more confident as I notice little things...So I felt like two things that really needed to happen before I felt really comfortable using the men’s restroom were the boobs had to go, the voice had to change, and the facial hair. And these would be ... when I had these three things I felt more safe...” | “If somebody were to really hate transgender people and they saw me, they could hurt me. And so the binder helps with that and also it helps my self-confidence build and also makes me feel normal.” |

### Themes

#### Physical health impacts

Among BTC participants, the most reported health conditions attributed to binding were back pain (n=20, 45%), shortness of breath (n=18, 42%), bad posture (n=12, 28%), and shoulder pain (n=12, 27%). For transmasculine participants who did not bind, no physical health problems were reported. The majority of transmasculine participants who bound expressed concern about the effects of binding on their health (n=25, 69%). Particularly, more than half of binding participants who used binders (69%), sports bras (55%), or shirt layering (54%) experienced back pain.

Participants in the TOP study experienced physical health impacts when binding, with duration of binding and binding materials mentioned as major factors. Physical challenges ranged from having full body itches to experiencing restriction while eating.

In addition, participants discussed the physical impacts of wearing binding material that were not breathable. However, participants emphasized binding was a necessity, despite the physical challenges. It is important for providers and caretakers to understand that although these physical challenges are painful, the benefits of binding outweigh them. This should be central to the way that binding discussions are approached by health providers and care takers.

### Provider de-prioritization of binding

Among the BTC participants, those who bound tried and were unable to access transition related care in the past 12 months at a higher frequency (81%) than those who did not bind (29%) (**Table 1**). Transmasculine survey participants reported discrimination by healthcare providers. 39% (n=17) of participants felt that healthcare providers were not comfortable with transgender people. Notably, 25% (n=11) had a healthcare professional try to stop them from being transgender and 18% (n=8) had experienced disrespect from healthcare providers in the past and reported this as a barrier to accessing care.

TOP participants described the lack of education on the importance and role of binding by their providers. Advising participants to not bind led to frustrating experiences. Participants described misunderstanding doctors disregarding any potential benefits of binding, leading them to advocate for themselves, explaining that binding was necessary for them to continue living.

Transmasculine participants who bound encountered insensitivity and inexperience from providers who instructed them to stop binding all together without realizing its importance. Providers who do not provide a welcoming environment to discuss binding practices only create additional barriers to optimal health care for transmasculine individuals who bind.

An additional barrier to care for TOP participants was financial costs to accessing safe binding materials. There is an urgent need for health care providers to prioritize and understand the importance of chest binding in their practices as an essential and potentially life-saving role. Patients who bind need support in accessing binding materials through their provider’s advocacy to connect them to these life-saving resources.

### Gender affirmation as a facilitator of safety

Among BTC participants, 86% of participants noted that being perceived as cisgender in public was important. Additionally, 52% of binding participants had a high score on the gender identity non-affirmation scale, where respondents answer questions related to being seen, understood, and respected as their gender (scale 0-20, high score > 10).^16^

Top participants described binding as a method of gender affirmation that ultimately preserved their safety. By binding, they also felt more comfortable by how they were perceived through societal norms. Participants explained how binding allows them to be more at ease in social situations. Binding can lead to both social and gender affirmation, as participants feel an increased sense of safety.

Further detailing their sense of safety while binding, TOP participants felt that binding helped them from experiencing microaggressions and discrimination. Participants described being less likely to be questioned for their gender, and therefore avoiding intrusive, and potentially discriminatory experiences that can compromise their safety. Binding can be a useful practice as a layer of protection from stigmatization against transgender individuals.

### The need for safer binding practice education

Most BTC participants bound in their lifetime (n=36, 82%), though fewer were binding at the time of the survey (n=13, 30%). The average age of binding initiation among those who bound was 28 years, ranging from 17 to 51 years. When binding, nearly half 43% (n=19) reported binding 7 days per week and 52% (n=23) bound 8+ hours per day on average. Most participants bound for 1 to 3 years total, suggesting that long-term binding is a common practice. Participants who used ace bandages or duct tape for binding named financial concerns as a barrier to accessing health care and the length of time they bound was from 1 to over 7 years.

Transmasculine TOP participants discussed the urgent need for access to safer binding practice education, especially when they began binding. Participants began their binding practice with ace bandages, and not learning about binders until later in their lives. Those interested in binding should be guided to binding resources so that they can practice binding in a way that reduces physical challenges.

TOP participants described their high binding frequency, detailing how binding is an everyday practice. Binding every day for long hours of time is outside of the clinical recommendations for binding, further emphasizing the importance of access to safer binding practices. As binding is a long-term practice for many, having resources about binding is essential to avoid long term physical health impacts such as back pain, skin irritation, fractured ribs, and infection.^3,17–19^ One long term health impact-reduced skin elasticity can impact surgical results among those who seek chest reconstruction surgery.^19^ If participants had access to information on safer binding practices and materials, they could have avoided major physical health impacts.

### Binding as mental health promotion

Among BTC participants, thirty-two percent (n=14) of transmasculine participants had PTSD symptoms in the past 30 days, 34% (n=15) had depressive symptoms (past 2 weeks), and 41% (n=18) had anxiety symptoms in the past 2 weeks.^20,21(p7),22(p2)^ There were no major differences in mental health outcomes by binding group, although our sample size is too small to test statistical significance (Table 1).

Binding led TOP participants closer to their gender affirmation goals, which led to improvements in their mental health. Participants described the central importance of binding on their internal sense of self and everyday experiences. By binding, participants mentioned being able to feel an improved sense of confidence. Binding should be understood as a mechanism to support and potentially improve one’s mental health.

Binding was especially important in social situations, as it helped build self-esteem. Participants alluded to the major impact binding has on feeling more at ease socially. Mental health improvement is an additional benefit of binding that providers should be informed about and incorporate into their practices.

## Discussion

Binding is an important part of gender affirmation for transmasculine people, with social and mental health benefits. While quantitative findings on time spent binding suggest that binding is a common practice, participants experienced notable physical impacts. The common use of binding and its potential physical health effects among BTC participants align with findings from the Binding Health Project results.^7^ Nearly half of transmasculine people bound for 7 days per week and 52% of participants bound for more than the clinically recommended limit of 8 hours per day.^23^ These findings along with the qualitative theme of physical impacts of pain indicate that there is need for improved provider education, safer binding support for transmasculine people who bind, and accessible pathways to top surgery for those interested.

While there may be negative physical health effects of binding from quantitative findings, qualitative findings suggested mental health benefits of reaching gender affirmation goals outweigh them. Providers should focus on reducing negative physical health effects by encouraging safe binding practices. The juxtaposition of choosing mental well-being over physical discomfort is portrayed in a 2019 Australian qualitative study of in-depth semi-structured interviews, where transmasculine participants thought of binding as a negotiation.^44^ Binding as mental health promotion is exemplified in a 2017 community-based survey of transmasculine people that found that binding was associated with reduced anxiety, depression, and dysphoria.^7,24^

Additionally, binding led to gender affirmation, which served as a facilitator of safety. For some, feeling safe largely depends on being perceived as cisgender (one’s gender aligning with sex-assigned-at-birth), and this process may include binding.^24^ A 2019 qualitative study documented how transmasculine participants considered binding as an important mechanism of controlling public perceptions.^4^

The diverse physical impacts of binding reported in our study speaks to the larger issue of access to affordable chest reconstruction surgery for transmasculine people who want this procedure. An online survey of 81 chest surgery patients reported significant improvement in their quality of life and reduced gender dysphoria related health outcomes in 86% of transmasculine patients.^25,26^ There may also be potential differences in access to chest surgery by socioeconomic status and race within transmasculine populations. Healthcare providers must understand these health disparities and barriers to care so they can advocate for access to this procedure, which has shown to alleviate mental health symptoms.^25,26^

BTC study participants reported that they felt healthcare providers were not comfortable with transgender people,which aligns with Binding Health Project results—more than half of transmasculine participants reported a healthcare provider reacted negatively to their binding.^7^ In the 2015 United States Transgender survey, one-third of participants who saw a healthcare provider experienced at least one negative encounter against their transgender identity, illustrating the need for more competency from health providers.^6^ Viewing binding practices as harmful to the patient creates an additional stigmatizing barrier to access safe binding practice information, and lack of support can delay healthcare seeking. This prevents transgender individuals from receiving medically necessary services for their gender transition, leading to subsequent poor quality of life and mental health.^23,27–29^

Transgender people often only have access to transgender-competent providers if they have access to larger transgender social networks, who share community recommendations by word of mouth.^28^ Most resources on binding health are not included in medical education curricula.^23,30,31^ Negative experiences with healthcare providers among BTC participants indicate an urgent need for more clinical health education related to chest binding—based on the standards of care for transgender people on binding practices and the growing body of peer-reviewed research on binding experiences.^23^

### Implications

Clinicians should be prepared to support transmasculine patients who bind and familiar with potential physical and mental health impacts.^5^ Recommendations for binding practices from community-based organizations and recent studies are accessible for providers and patients.^2–4,7,17,28,32,33^ Resources for accessing affordable and safer binders should be well understood.

Healthcare providers must create transgender-competent environments to discuss safe binding practices to prevent long-term health consequences and prioritize the mental and social benefits of this practice. With threats to the rights of trans youth across the United States, it is essential for healthcare providers to provide affirming binding health support, especially in states where gender affirming services are limited or prohibited.^34^ Health policy makers should consider healthcare coverage of binders. This could reduce barriers to care, improve mental and physical health, and lead to more referrals to gender affirming care if desired.

### Conclusion

Binding is an important part of gender affirmation for many transmasculine people, with social and mental health benefits. However, many transmasculine people report barriers to social and healthcare provider support around binding. Healthcare providers should understand the benefits of binding and be prepared to discuss safer binding practices with transmasculine patients.

### Limitations

This study is not without limitations. The cross-sectional data from the BTC study limits our understanding of temporality between binding practices and their effects. Convenience sampling limits the generalizability of this study, and the small sample size limits complex statistical analyses. Most participants were White, had access to higher education, and were insured, potentially limiting transferability of the data.

## Data Availability

N/A

## Acknowledgements

The authors would like to thank the participants from the BTC and TOP studies for their participation, time, and contribution to transgender health research.

## Ethical Approval

Protocols and procedures for both studies were approved by the Johns Hopkins School of Public Health Institutional Review Board.

## Abbreviations Used

BTC: Be the Conversation

LGBTQ: Lesbian Gay Bisexual Transgender Queer

## References

1. Olson-Kennedy J, Warus J, Okonta V, Belzer M, Clark LF. Chest Reconstruction and Chest Dysphoria in Transmasculine Minors and Young Adults. JAMA Pediatr. 2018;172(5):431–436. doi:10.1001/jamapediatrics.2017.5440

2. Julian JM, Salvetti B, Held JI, Murray PM, Lara-Rojas L, Olson-Kennedy J. The Impact of Chest Binding in Transgender and Gender Diverse Youth and Young Adults. J Adolesc Health. Published online October 27, 2020. doi:10.1016/j.jadohealth.2020.09.029

3. Hudson. Hudson’s Guide: FTM Binding. Published 2004. Accessed January 27, 2022. http://www.ftmguide.org/binding.html

4. Lee A, Simpson P, Haire B. The binding practices of transgender and gender-diverse adults in Sydney, Australia. Cult Health Sex. 2019;21(9):969–984. doi:10.1080/13691058.2018.1529335

5. Reisner SL, Gamarel KE, Dunham E, Hopwood R, Hwahng S. Female-to-Male Transmasculine Adult Health: A Mixed-Methods Community-Based Needs Assessment. J Am Psychiatr Nurses Assoc. 2013;19(5):293–303. doi:10.1177/1078390313500693

6. James SE, Herman J, Keisling M, Mottet L, Anafi M. 2015 U.S. Transgender Survey (USTS): Version 1. Published online 2019. doi:10.3886/ICPSR37229.V1

7. Peitzmeier S, Gardner I, Weinand J, Corbet A, Acevedo K. Health impact of chest binding among transgender adults: a community-engaged, cross-sectional study. Cult Health Sex. 2017;19(1):64–75. doi:10.1080/13691058.2016.1191675

8. Factor R, Rothblum E. Exploring gender identity and community among three groups of transgender individuals in the United States: MTSs, FTMs, and genderqueers. Health Sociol Rev. 2008;17(3):235–253. doi:10.5172/hesr.451.17.3.235

9. Bockting WO, Knudson G, Goldberg JM. Counseling and Mental Health Care for Transgender Adults and Loved Ones. Int J Transgenderism. 2006;9(3-4):35–82. doi:10.1300/J485v09n03_03

10. Marinkovic M, Newfield RS. Chest reconstructive surgeries in transmasculine youth: Experience from one pediatric center. Int J Transgenderism. 2017;18(4):376–381. doi:10.1080/15532739.2017.1349706

11. Creswell JW. Research Design: Qualitative, Quantitative, and Mixed Methods Approaches. 4th ed. SAGE Publications; 2014.

12. Vaismoradi M, Turunen H, Bondas T. Content analysis and thematic analysis: Implications for conducting a qualitative descriptive study. Nurs Health Sci. 2013;15(3):398–405. doi:10.1111/nhs.12048

13. Kiperman S, Schacter HL, Judge M, DeLong G. LGBTQ+ Youth’s Identity Development in the Context of Peer Victimization: A Mixed Methods Investigation. Int J Environ Res Public Health. 2022;19(7):3921. doi:10.3390/ijerph19073921

14. Guetterman TC, Fetters MD, Creswell JW. Integrating Quantitative and Qualitative Results in Health Science Mixed Methods Research Through Joint Displays. Ann Fam Med. 2015;13(6):554–561. doi:10.1370/afm.1865

15. Creswell JW, Clark VLP. Designing and Conducting Mixed Methods Research. Sage Publications, Inc; 2007:xviii, 275.

16. Testa RJ, Habarth J, Peta J, Balsam K, Bockting W. Development of the Gender Minority Stress and Resilience Measure. Psychol Sex Orientat Gend Divers. 2015;2(1):65–77. doi:10.1037/sgd0000081

17. Chest Binding 101 - FTM Binder Guide | FTM Binding, Chest Binder, Breast Binders. TransGuys.com. Published September 7, 2010. Accessed January 27, 2022. http://transguys.com/features/chest-binding

18. Monstrey S, Selvaggi G, Ceulemans P, et al. Chest-wall contouring surgery in female-to-male transsexuals: a new algorithm. Plast Reconstr Surg. 2008;121(3):849–859. doi:10.1097/01.prs.0000299921.15447.b2

19. Berry MG, Curtis R, Davies D. Female-to-male transgender chest reconstruction: a large consecutive, single-surgeon experience. J Plast Reconstr Aesthetic Surg JPRAS. 2012;65(6):711–719. doi:10.1016/j.bjps.2011.11.053

20. Primary Care PTSD Screen for DSM-5 (PC-PTSD-5) - PTSD: National Center for PTSD. Accessed April 4, 2022. https://www.ptsd.va.gov/professional/assessment/screens/pc-ptsd.asp

21. Williams N. The GAD-7 Questionnaire. Occup Med. 2014;64(3):224–224. doi:10.1093/occmed/kqt161

22. Patient Health Questionnaire (PHQ-9 & PHQ-2). https://www.apa.org. Accessed April 4, 2022. https://www.apa.org/pi/about/publications/caregivers/practice-settings/assessment/tools/patient-health

23. Standards of Care for the Health of Transsexual, Transgender, and Gender Nonconforming People [7th Version]. Published online 2012. Accessed January 27, 2022. https://www.wpath.org/publications/soc

24. Couch M, Pitts M, Mulcare H, Croy S, Mitchell A, Patel S. TranZnation - a Report on the Health and Wellbeing of Transgendered People in Australia and New Zealand. Australian Research Centre in Sex, Health and Society; 2007. Accessed May 18, 2021. https://apo.org.au/node/3849

25. Davis SA, Meier SC. Effects of Testosterone Treatment and Chest Reconstruction Surgery on Mental Health and Sexuality in Female-To-Male Transgender People. Int J Sex Health. 2014;26(2):113–128. doi:10.1080/19317611.2013.833152

26. Poudrier G, Nolan IT, Cook TE, et al. Assessing Quality of Life and Patient-Reported Satisfaction with Masculinizing Top Surgery: A Mixed-Methods Descriptive Survey Study. Plast Reconstr Surg. 2019;143(1):272–279. doi:10.1097/PRS.0000000000005113

27. Poteat T, German D, Kerrigan D. Managing uncertainty: a grounded theory of stigma in transgender health care encounters. Soc Sci Med 1982. 2013;84:22–29. doi:10.1016/j.socscimed.2013.02.019

28. A health chest resource for trans folk. QMUNITY. Accessed February 2, 2021. https://qmunity.ca/resources/i-heart-my-chest/

29. Cruz TM. Assessing access to care for transgender and gender nonconforming people: a consideration of diversity in combating discrimination. Soc Sci Med 1982. 2014;110:65–73. doi:10.1016/j.socscimed.2014.03.032

30. Coleman E, Bockting W, Botzer M, et al. Standards of Care for the Health of Transsexual, Transgender, and Gender-Nonconforming People, Version 7. Int J Transgenderism. 2012;13(4):165-232. doi:10.1080/15532739.2011.700873

31. Herman H. Physical Therapy and Gender Affirmation. Accessed February 12, 2021. https://fenwayhealth.org/wp-content/uploads/15b.-Physical-Therapy.pdf

32. Jarrett BA, Corbet AL, Gardner IH, Weinand JD, Peitzmeier SM. Chest Binding and Care Seeking Among Transmasculine Adults: A Cross-Sectional Study. Transgender Health. 2018;3(1):170–178. doi:10.1089/trgh.2018.0017

33. Binding, Packing, Tucking & Padding. Accessed November 14, 2021. http://www.phsa.ca/transcarebc/care-support/transitioning/bind-pack-tuck-pad

34. Conron KJ, O’Neill K, Vasquez L, Mallory C. Prohibiting Gender-Affirming Medical Care for Youth. Published online March 2022. Accessed April 5, 2022. https://williamsinstitute.law.ucla.edu/publications/bans-trans-youth-health-care/

